# Targeted Analysis of Whole Exome Sequencing Data Identifies Biologically Plausible Causal Variants in Genetically Unresolved Oculocutaneous Albinism Patients From India

**DOI:** 10.64898/2026.09.03.26362137

**Authors:** Tithi Dutta, Debrup Dey, Arpan Saha, Santasree Banerjee, Kunal Ray, Mainak Sengupta

## Abstract

**Background:** Oculocutaneous albinism (OCA) is a genetically heterogeneous disorder characterized by hypopigmentation and visual abnormalities. Despite eight established OCA-associated loci, some clinically diagnosed patients remain genetically unresolved, suggesting contributions from additional pigmentation-associated genes.

**Methods:** Whole exome sequencing was performed in clinically diagnosed OCA patients lacking pathogenic variants or carrying a single heterozygous variant in known OCA genes (OCA1–OCA8). Targeted analysis of 809 pigmentation-associated genes involved in melanogenesis, melanosomal transport, melanocyte differentiation, stem-cell maintenance, and pigmentation phenotypes was conducted. Variants were prioritized based on ACMG classification, rarity, predicted coding consequence, and biological relevance to pigmentation pathways.

**Results:** In 5 patients with missing heritability, ACMG/Varsome/Ensembl Variant Effect Predictor-classified pathogenic/likely pathogenic rare variants were identified in pigmentation-associated genes including *RAB38, MYO5A*, and *DOCK7*, involved in melanosome biogenesis/maturation, transport, or pigmentation abnormalities. Potential disease-causing variants were also identified in *BMPR1B, MYC, POLG, GNA11*, and *GGT1*, which lack established roles in melanin biosynthesis or distribution but may contribute to pigmentation based on emerging evidence. Two patients harboured prioritised variants in multiple pigmentation-associated genes. Notably, 3 of 5 patients harboured previously identified changes in known OCA-causing genes in heterozygous condition, suggesting cumulative modifier or digenic/oligogenic contributions.

**Conclusions:** Our findings broaden the spectrum of candidate pigmentation-associated variants in unresolved OCA cases and support possible oligogenic or modifier-driven mechanisms underlying phenotypic heterogeneity. However, these findings are hypothesis-generating and require functional validation.

## Introduction

Oculocutaneous albinism (OCA) comprises a heterogeneous group of inherited disorders characterized by reduced or absent melanin synthesis, thus affecting the skin, hair, and eyes. Clinically, OCA is associated with hypopigmentation, nystagmus, reduced visual acuity, foveal hypoplasia, and photophobia. To date, pathogenic variants in 8 loci and 7 underlying genes viz. *TYR, OCA2, TYRP1, SLC45A2, SLC24A5, LRMDA*, and *DCT* have been implicated in nonsyndromic OCA (Dutta et al., 2024; Zaman et al., 2024).

Despite advances in sequencing techniques, ∼50% of clinically diagnosed OCA patients remain genetically unresolved or only partially explained by a single heterozygous variant in a known OCA-associated gene ^5–7^ Increasing evidence indicates that additional pigmentation-associated genes may contribute to disease manifestation through modifier effects, or putative digenic/oligogenic inheritance ^3,14,16,20^

In the present study, whole exome sequencing followed by targeted analysis of pigmentation-associated genes beyond the known OCA-causing genes was performed in genetically unresolved OCA patients to address the issue of missing heritability in OCA.

## Materials and Methods

Whole exome sequencing was performed in 14 clinically diagnosed OCA patients from different parts of India, who either lacked any pathogenic variants or harboured only a single heterozygous variant in known OCA-associated genes (OCA1–OCA8) (Mondal et al., 2012; Sengupta et al., 2007, 2010, unpublished data).

A curated gene panel comprising 809 genes was generated for variant prioritization. The panel included (i) human orthologues of murine genes reported to influence coat-colour phenotypes in the Color Genes dataset [http://www.espcr.org/micemut] (ii) all known genes implicated in ocular albinism (OA), and syndromic forms of albinism if they are already not represented within the murine orthologue dataset, and (iii) genes identified as targets through genome-wide association studies (GWAS) with pigmentation-related traits. Collectively, the selected set of genes were involved in pigmentation disorders, melanogenesis, melanosome biogenesis and transport, melanocyte differentiation, and melanocyte stem-cell biology.

Rare coding variants with a global minor allele frequency ≤0.01 and classified as Pathogenic, Likely Pathogenic, Pathogenic Moderate, or Pathogenic Supporting according to the American College of Medical Genetics and Genomics (ACMG) guidelines or VarSome (v13.17.1.0) or the Ensembl Variant Effect Predictor (VEP) were prioritised as potential disease-causing mutations.

To evaluate the potential structural consequences of the prioritized variants, protein structures of the concerned genes were retrieved from the AlphaFold Protein Structure database and assessed using DynaMut2.=. The amino acid sequences were retrieved from UniProt. The change in thermodynamic stability of a protein harbouring the mutation was calculated in terms of ΔΔG (ΔΔG= ΔGwild-typeΔGMutant), where ΔG is the difference between the free energies of the folded and unfolded states, and a mutation with ΔΔG < 0 indicates the reduced stabilization of protein structure.

Owing to limited availability of archived samples and the absence of parental DNA, segregation analysis and Sanger sequencing validation could not be performed for any of the samples. Therefore, to overcome the burden of false positives, novel variants lacking prior documentation were excluded from the current study, and only previously reported variants with assigned rsIDs were considered for downstream analyses.

The study was designed as an exploratory analysis of biologically plausible pigmentation-associated variants in unresolved OCA cases.

## Results

Targeted analysis of whole exome sequencing data prioritized variants in 5 genetically unresolved OCA patients.

<u>Patient OC115</u>, previously identified with a heterozygous mutation in the OCA 8-associated gene *DCT* (*unpublished data*), was found to harbor the heterozygous variant NM_022337.3:c.199G>T NP_071732.1:(p.Ala67Pro) in *RAB38*. The patient was additionally found to carry *MYC:* NM_002467.6: c.1091A>C NP_002458.2:p. Asp364Val; *POLG:* NM_002693.3:c.452T>C NP_001119603.1:p.Leu151Gln; and *GNA11*: NM_002067.5:c.1023C>A NP_002058.2: (p.Phe341Leu), all in heterozygous condition.

<u>Patient OC51</u>, previously identified with a heterozygous mutation in the OCA 1-associated gene *TYR* (Chaki et al.,2011) carried a homozygous *DOCK7* variant: NM_001367561.1: c.281G>A NP_001354490.1: (p.Arg94Gln).

<u>Patient OC21</u> harbored a heterozygous *MYO5A* variant NM_001382347.1: c.2692C>T NP_001369276.1: (p.Arg898Trp).

<u>Patients OC5 and OC15</u> carried rare *GGT1* variants: NM_001288833.2: c.1063C>T, NP_001275762. 1:(p.Arg355Trp) in both the patients; and NM_001288833.2: c.1162G>A, NP_001275762. 1:(p.Ala388Thr) exclusively in OC15. OC5 incidentally also harbored a *BMPR1B* homozygous variant NM_001203.3:c.1385G>A, NP_001194.1: (p.Cys462Tyr) in the backdrop of a previously identified splice-site mutation in the OCA 4-associated gene *SLC45A2* (Sengupta et al.,2007).

A summary of the identified variants and the associated genes is provided in Table 1.

**Table 1.** Variants prioritised from targeted analysis of whole exome sequencing data for genetically unresolved OCA patients.

| ID | Ethnicity | Known status of OCA genes (I-VIII) | Prioritised variants identified |
| --- | --- | --- | --- |
| OC115 | Madiga | **DCT:** rs759890417 p.Gly236Ala (Hh) | **GNA11:** rs140749796; p.Phe341Leu (Hh);<br>**MYC:** rs746333391; p.Asp364Val (Hh);<br>**POLG:** rs749018627; p.Leu151Gln (Hh);<br>**RAB38:** rs777613432; p.Ala67Pro (Hh) |
| OC21 | Paadmasali | NA | **MYO5A:** rs754463537 p.Arg898Trp (Hh) |
| OC51 | Vysyas | **TYR:** rs747995722 p.Glu219Lys (Hh) | **DOCK7:** rs570514826. p.Arg94Gln (HH) |
| OC5 | Reddi | **SLC45A2:** c.1032+1G>A (Hh) | **GGT1:** rs200419006; p.Arg355Trp (Hh);<br>**BMPR1B:** rs766596192; p, Cys462Tyr (HH) |
| OC15 | Perika | NA | **GGT1:** rs200419006; p.Arg355Trp (Hh);<br>rs761538704; p.Ala388Thr (Hh) |

*In silico* structural analysis revealed that all the alternate mutant alleles were predicted to reduce the structural stability of the corresponding proteins, except for the *MYO5A* variant NM_001382347.1:c.2692C>T, NP_001369276.1: (p.Arg898Trp), which exhibited a stabilizing effect. The detailed results of the structural stability and steric clash analyses are shown **(SF-IA and SF-IB)**

## Discussion

The present study identified rare pathogenic/likely pathogenic variants in multiple biologically relevant pigmentation-associated genes beyond the known OCA-causing genes, in clinically diagnosed patients with unresolved heritability. The following section discusses the biological plausibility of these genes to contribute to hypopigmentation in such patients. RAB38 is a melanocyte-enriched Rab GTPase that controls a critical trafficking step in melanosome biogenesis, i.e the delivery of tyrosinase (TYR) and TYRP1 from the trans-Golgi network (TGN) to immature melanosomes. In Rab38/Rab32-deficient melanocytes, TYR is mistargeted after TGN exit and undergoes enhanced lysosomal degradation, leading to reduced melanin synthesis and hypopigmentation. ^21^ Consistent with this mechanism, loss-of-function mutations in RAB38 cause hypopigmentation in multiple animal models, including mice. ^9^

GNA11 participates in melanocyte differentiation pathways and has been associated with altered pigmentation phenotypes in murine models. ^13^ Unlike canonical OCA genes that directly impair melanin synthesis or melanosome biogenesis, GNA11 functions upstream by regulating the transcriptional competence of melanocytes, including MITF-dependent expression of TYR and other pigmentation genes. Thus, hypomorphic or dysregulated GNA11 signalling could reduce the expression or functional maturation of melanogenic enzymes, acting as a modifier that amplifies the phenotypic impact of a TYR variant. ^19^ MYC is a master transcriptional regulator of cell proliferation that contributes to melanocyte stem cell (McSC) maintenance and expansion in the hair follicle niche. Disruption of MYC-associated pathways in murine models results in progressive coat hypopigmentation, ^10^ consistent with impaired McSC proliferation or premature stem cell depletion. Reduced MYC activity could therefore diminish the pool of differentiated melanocytes, lower total tissue TYR enzyme and unmask a subclinical TYR defect.

POLG may indirectly influence pigmentation through mitochondrial dysfunction, oxidative stress, and impaired melanocyte stem-cell survival.^1,23^ Therefore, POLG-associated mitochondrial stress may act as a modifier by reducing melanocyte numbers and/or impairing the function of existing melanocytes, thereby exacerbating the phenotypic expression of a TYR variant.

*DOCK*_7_ encodes a Rho-family guanine nucleotide exchange factor that regulates actin cytoskeleton dynamics and cell migration. Dock_7_ mutant mice exhibit generalized hypopigmentation and white spotting due to defective melanoblast migration during development and impaired melanocyte maintenance in postnatal tissues.^2^ Although DOCK_7_ has not yet been established as a cause of human pigmentation disorders, its role in melanoblast positioning and melanocyte persistence renders it a biologically plausible modifier. We hypothesise that concurrent TYR heterozygous and DOCK_7_ homozygous variants may synergistically reduce melanocyte density and total TYR activity, contributing to the observed OCA-like phenotype.

*MYO5A* encodes myosin-Va, an actin-based motor protein essential for melanosome transport along dendritic actin filaments. Pathogenic MYO5A variants cause Griscelli syndrome type 1, characterized by pigmentary dilution due to perinuclear melanosome aggregation, while murine Myo5a mutants exhibit coat-colour dilution ^15^.When combined with a TYR variant that reduces melanin synthesis, defective melanosome trafficking could compound hypopigmentation, yielding an OCA-like phenotype

BMPR1B participates in BMP signalling, which has recognised roles in neural crest development and melanocyte differentiation. Although BMPR1B has not been implicated directly in OCA, perturbation of BMP signalling may influence melanocyte development and survival, rendering it a plausible modifier candidate. ^8^

*GGT1* encodes gamma-glutamyl transferase 1, which participates in glutathione metabolism and maintenance of intracellular redox homeostasis, and thus, is posed as a biologically plausible modifier of pigmentation, particularly through regulating oxidative stress pathways. Oxidative balance is important for melanocyte survival and melanogenesis, and disruption of cellular redox mechanisms may theoretically influence pigmentation phenotypes through indirect modifier effects. ^4^ In fact, glutathione levels influence the balance between eumelanin and pheomelanin. ^11^

An important observation from the present study is that 3 patients harboured a heterozygous pathogenic variant in a recognised OCA-associated gene together with one or more potentially deleterious variants in additional pigmentation-related genes. Although none of these secondary variants can independently explain the phenotype, their coexistence raises the possibility of an oligogenic model of inheritance. In such a scenario, a heterozygous variant in an established OCA gene may create a state of partial melanogenic insufficiency, while additional variants affecting melanosome biogenesis, intracellular trafficking, melanocyte differentiation, redox homeostasis, or related pathways collectively reduce pigment production below a critical functional threshold. Such cumulative effects have increasingly been proposed to explain missing heritability and variable expressivity in Mendelian disorders. While the present data do not establish causality, they are exploratory findings that support the hypothesis that unresolved OCA cases may, in some instances, result from the combined contribution of multiple hypomorphic alleles rather than a single highly penetrant pathogenic variant.

The study has several limitations. Segregation analysis and functional validation could not be performed because of limited archived sample availability and lack of parental DNA. Consequently, the pathogenic contribution of the identified variants cannot be conclusively established. Furthermore, the identified variants have previously been reported in population databases without established association with albinism phenotypes. Nevertheless, the present findings broaden the spectrum of candidate pigmentation-associated variants identified in unresolved OCA patients and underscore the importance of considering modifier-based and oligogenic mechanisms in pigmentation disorders. Then again, the study was exploratory and intended to identify biologically plausible candidate modifiers rather than establish any statistical enrichment. These should therefore be considered hypothesis-generating and require further validation in larger cohorts.

## Conclusions

The identification of rare pathogenic or likely pathogenic variants in multiple biologically plausible pigmentation-associated genes in genetically unresolved OCA patients in the background of a heterozygous mutation in a canonical OCA gene raises the possibility of modifier effects or oligogenic inheritance contributing to disease expression. This observation aligns with the broader concept of missing heritability in Mendelian disorders, where phenotypic severity may reflect the cumulative burden of variants across multiple loci rather than a single causative mutation. Such polygenic or oligogenic architectures could explain variable penetrance and phenotypic heterogeneity among patients with possible similar primary mutations. However, segregation analyses in larger cohorts, coupled with functional validation, are required to establish the pathogenic relevance of these candidate modifier variants and to define their contribution to the overall genetic architecture of OCA-like phenotypes.

## Data Availability

All data produced in the present work are contained in the manuscript

## Acknowledgements

We would like to thank the DST-SERB for providing funds for the completion of the project (CRG/2019/001487), Department of Science and Technology-Promotion of University Research and Scientific Excellence [DST-PURSE], Government of India for providing funds to the University of Calcutta for academic infrastructural facilities. T. Dutta and A.Saha was supported by Senior Research Fellowship from University Grant Commission [UGC], Government of India. We have not received any extramural funding for the preparation of data or the manuscript.

## Author contributions

T. Dutta, and M. Sengupta conceptualized and designed the study. D.Dey and A.Saha contributed to the literature and database searches and data curation. T.Dutta, M.Sengupta, A.Saha contributed to the data arrangement. T. Dutta drafted the manuscript with important intellectual contribution from M. Sengupta. S. Banerjee helped in analysis of data, provoded substantial intellectual inputs and critically reviewed the manuscript; K.Ray contributed to the collection of the patient samples; and M. Sengupta critically revised the manuscript, figures, tables, and supplementary data and supervised the entire work. All authors provided critical feedback and helped in shaping the research, analysis, and manuscript. None of the authors has received extramural funding for the preparation of data or the manuscript.

## Ethics approval

Any aspect of the work covered in this manuscript that has involved either experimental animals or human patients has been conducted with the ethical approval of all relevant bodies and that such approvals are acknowledged within the manuscript.

## Data availability

All data generated or analysed during this study are included in this article and its supplementary information files.

## Supplementary Files (SF)

SF-IA: The result table showing the pathogenicity of the variants and its effect on the protein structure (ΔΔG value) and alteration of steric clash for wild type and mutant variant.

SF-IB: The protein structural representation of the wild type and mutant variant obtained from DynaMut2 and AlphaFold. (ppt)

## Reference

Baxter, L. L., Watkins-Chow, D. E., Pavan, W. J., & Loftus, S. K. (2019). A curated gene list for expanding the horizons of pigmentation biology. In Pigment Cell and Melanoma Research (Vol. 32, Number 3, pp. 348–358). Blackwell Publishing Ltd. 10.1111/pcmr.12743

Blasius, A. L., Brandl, K., Crozat, K., Xia, Y., Khovananth, K., Krebs, P., Smart, N. G., Zampolli, A., Ruggeri, Z. M., & Beutler, B. A. (2009). Mice with mutations of Dock7 have generalized hypopigmentation and white-spotting but show normal neurological function. Proceedings of the National Academy of Sciences of the United States of America, 106(8), 2706–2711. 10.1073/pnas.0813208106

Butkovič, R., Healy, M. D., de Heus, C., Walker, A. P., Beyers, W., McNally, K. E., Lewis, P. A., Heesom, K. J., Liv, N., Klumperman, J., Di Pietro, S., Collins, B. M., & Cullen, P. J. (2025). Identification of a RAB32-LRMDA-Commander membrane trafficking complex reveals the molecular mechanism of human oculocutaneous albinism type 7. Nature Communications, 16(1). 10.1038/s41467-025-63855-8

Chaubal, V. A., Nair, S. S., Ito, S., Wakamatsu, K., & Mojamdar, M. V. (2002). γ-glutamyl transpeptidase and its role in melanogenesis: Redox reactions and regulation of tyrosinase. Pigment Cell Research, 15(6), 420–425. 10.1034/j.1600-0749.2002.02004.x

Dutta, T., Ganguly, K., Saha, A., Sil, A., Ray, K., & Sengupta, M. (2024). Identifying genetic defects in oculocutaneous albinism patients of West Bengal, Eastern India. Molecular Biology Reports, 51(1). 10.1007/s11033-024-09777-y

Ganguly, K., Dutta, T., Saha, A., Sarkar, D., Sil, A., Ray, K., & Sengupta, M. (2020). Mapping the TYR gene reveals novel and previously reported variants in Eastern Indian patients highlighting preponderance of the same changes in multiple unrelated ethnicities. Annals of Human Genetics, 84(3), 303–312. 10.1111/ahg.12376

Green, D. J., Michaud, V., Lasseaux, E., Plaisant, C., Fitzgerald, T., Birney, E., Black, G. C., Arveiler, B., & Sergouniotis, P. I. (2024). The co-occurrence of genetic variants in the TYR and OCA2 genes confers susceptibility to albinism. Nature Communications, 15(1). 10.1038/s41467-024-52763-y

Infarinato, N. R., Stewart, K. S., Yang, Y., Gomez, N. C., Amalia Pasolli, H., Hidalgo, L., Polak, L., Carroll, T. S., & Fuchs, E. (2020). BMP signaling: At the gate between activated melanocyte stem cells and differentiation. Genes and Development, 34(23–24), 1713–1734. 10.1101/gad.340281.120

Kriebel, W. G., Larimer-Picciani, A. M., Nukala, M., Sahel, J. A., & Byrne, L. C. (2026). Quantifying functional vision in a mouse model of oculocutaneous albinism type 1. Scientific Reports, 16(1). 10.1038/s41598-026-45301-x

Lee, J. H., & Fisher, D. E. (2014). Melanocyte stem cells as potential therapeutics in skin disorders. In Expert Opinion on Biological Therapy (Vol. 14, Number 11, pp. 1569–1579). Informa Healthcare. 10.1517/14712598.2014.935331

Lu, Y., Tonissen, K. F., & Di Trapani, G. (2021). Modulating skin colour: Role of the thioredoxin and glutathione systems in regulating melanogenesis. In Bioscience Reports (Vol. 41, Number 5). Portland Press Ltd. 10.1042/BSR20210427

Mondal, M., Sengupta, M., Samanta, S., Sil, A., & Ray, K. (2012). Molecular basis of albinism in India: Evaluation of seven potential candidate genes and some new findings. Gene, 511(2), 470–474. 10.1016/j.gene.2012.09.012

Moore, A. R., Ran, L., Guan, Y., Sher, J. J., Hitchman, T. D., Zhang, J. Q., Hwang, C., Walzak, E. G., Shoushtari, A. N., Monette, S., Murali, R., Wiesner, T., Griewank, K. G., Chi, P., & Chen, Y. (2018). GNA11 Q209L Mouse Model Reveals RasGRP3 as an Essential Signaling Node in Uveal Melanoma. Cell Reports, 22(9), 2455–2468. 10.1016/j.celrep.2018.01.081

Nagy, N., Pal, M., Kun, J., Galik, B., Urban, P., Medvecz, M., Fabos, B., Neller, A., Abdolreza, A., Danis, J., Szabo, V., Yang, Z., Fenske, S., Biel, M., Gyenesei, A., Adam, E., & Szell, M. (2024). Missing Heritability in Albinism: Deep Characterization of a Hungarian Albinism Cohort Raises the Possibility of the Digenic Genetic Background of the Disease. International Journal of Molecular Sciences, 25(2). 10.3390/ijms25021271

Pan, J., Zhou, R., Yao, L.-L., Zhang, J., Zhang, N., Cao, Q.-J., Sun, S., & Li, X. (2024). Melanophilin mediates the association of myosin-5a with melanosome via three distinct interactions. 10.7554/eLife.93662.2

Sauermann, R., Fancourt, B., Faulkner, T., Shute, H., Reid, D., Pask, A. J., & Feigin, C. Y. (2025). Loss-of-function mutations in ASIP and MC1R are associated with coat colour variation in marsupials. Biology Letters, 21(10). 10.1098/rsbl.2025.0302

Sengupta, M., Chaki, M., Arti, N., & Ray, K. (2007). SLC45A2 variations in Indian oculocutaneous albinism patients. Molecular Vision, 13(June), 1406–1411.

Sengupta, M., Mondal, M., Jaiswal, P., Sinha, S., Chaki, M., Samanta, S., & Ray, K. (2010). Comprehensive analysis of the molecular basis of oculocutaneous albinism in Indian patients lacking a mutation in the tyrosinase gene. British Journal of Dermatology, 163(3), 487–494. 10.1111/j.1365-2133.2010.09830.x

Silva-Rodríguez, P., Fernández-Díaz, D., Bande, M., Pardo, M., Loidi, L., & Blanco-Teijeiro, M. J. (2022). GNAQ and GNA11 Genes: A Comprehensive Review on Oncogenesis, Prognosis and Therapeutic Opportunities in Uveal Melanoma. In Cancers (Vol. 14, Number 13). MDPI. 10.3390/cancers14133066

Tingaud-sequeira, A., Mercier, E., Michaud, V., Pinson, B., Gazova, I., Gontier, E., Decoeur, F., McKie, L., Jackson, I. J., Arveiler, B., & Javerzat, S. (2022). The Dct™/™ Mouse Model to Unravel Retinogenesis Misregulation in Patients with Albinism. Genes, 13(7). 10.3390/genes13071164

Wasmeier, C., Romao, M., Plowright, L., Bennett, D. C., Raposo, G., & Seabra, M. C. (2006). Rab38 and Rab32 control post-Golgi trafficking of melanogenic enzymes. Journal of Cell Biology, 175(2), 271–281. 10.1083/jcb.200606050

Zaman, Q., Khan, J., Ahmad, M., Khan, H., Chaudhary, H. T., Rehman, G., Rahman, O. U., Shah, M. M., Hussain, J., Jamal, Q., Khan, B. T., Khan, M. A., Sadeeda Sahar, K., Idrees, M., Ahmad, R., Faisal, M. S., Khan, M. I., Khisroon, M., … Naseer, M. I. (2024). Unveiling genetics of non-syndromic albinism using whole exome sequencing: A comprehensive study of TYR, TYRP1, OCA2 and MC1R genes in 17 families. Gene, 894. 10.1016/j.gene.2023.147986

Zhang, R., Li, T., Du, F., Huang, J., Yu, N., & Long, X. (2026). Mitochondrial Dynamics in Skin Health and Disease: Energy, Ageing, and Therapeutic Perspectives. Burns & Trauma. 10.1093/burnst/tkag008

